# Predicted serotype distribution in invasive pneumococcal disease (IPD) among children less than five years prior to the introduction of the Pneumococcal Conjugate Vaccine (PCV) in Nigeria

**DOI:** 10.1101/2022.03.11.22272265

**Authors:** Aishatu L Adamu, John. Ojal, Isa S. Abubakar, Musa M. Bello, Kofo Odeyemi, Christy A.N. Okoromah, Victor Inem, Boniface Karia, Angela Karani, Donald. Akech, Katherine Gallagher, J. Anthony G Scott, Ifedayo M.O. Adetifa

## Abstract

**Background:** The 10-valent pneumococcal conjugate vaccine (PCV10) was introduced in Nigeria without any baseline data on serotype distribution in invasive pneumococcal disease (IPD). To estimate the proportion of IPD attributable to different serotypes, in children aged <5 years, we used statistical models based on the serotype-specific nasopharyngeal carriage prevalence and invasive capacity (IC).

**Methods:** We used the carriage data from one urban and one rural setting in Nigeria, collected within five months of PCV10 introduction (2016). For Model A, we used serotype-specific adult case-fatality ratios from Denmark as proxy for IC. In the second model, we used the ratio of IPD proportions to carriage prevalence (case-carrier ratios) from Kenya (Model B) and the ratio of IPD incidence to carriage acquisition (attack rates) from the UK (Model C) as measures of serotype IC.

**Results:** The models predict that serotypes with high carriage prevalence (6A, 6B, 19F and 23F) will dominate IPD. Additionally, Models B and C predictions emphasize serotypes 1, 4, 5, and 14, which were not prevalent in carriage but had high IC estimates. Non-PCV10 serotypes,6A and 19A, also dominated IPD predictions across models and settings. The predicted proportion of IPD attributed to PCV10 serotypes varied between 56% and 74% by model and setting.

**Conclusion:** Carriage data can provide preliminary insights into IPD serotypes in settings that lack robust IPD data. The predicted PCV10-serotype coverage for IPD was moderately high. However, predictions for non-PCV10 serotypes indicate that higher-valency PCVs that cover serotypes 6A and 19A may have a larger impact on IPD reductions.

---

Globally, the pneumococcus was estimated to cause 14 million cases of pneumonia and Invasive Pneumococcal Disease (IPD) and 800,000 deaths among children aged <5 years (U5s) prior to wide uptake of Pneumococcal Conjugate Vaccine (PCV).(1) Ten countries contribute to more than two-thirds of global pneumococcal deaths among U5s, and Nigeria, which has the highest burden of pneumococcal disease in sub-Saharan Africa (sSA), is one of these countries.(1,2) The 100 pneumococcal serotypes differ in their patterns of nasopharyngeal (NP) acquisition, persistent colonization, and NP clearance and also in their capacity to cause IPD of varying severity.(3–6) The majority of IPD is caused by a small proportion of these serotypes, and this distribution largely guided the choice of serotypes included in the licensed PCVs.(7) However, variations in relative serotype distribution in IPD have also been documented by geographic location and age.(7,8) For instance, recognition of the relative importance of serotypes 1 and 5, particularly in low and middle-income countries (LMICs), supported their inclusion in the 10-valent (PCV10) and 13-valent (PCV13) vaccines to replace the 7-valent PCV (PCV7).(7,9) The Serum Institute of India’s new 10-valent PCV (SII-PCV) substitutes serotype 4 and 18C in PCV10 with serotypes 6A and 19A to improve vaccine serotype coverage in LMICs.(10). Introduction of PCV to childhood vaccine programs has successfully reduced the burden of IPD caused by serotypes included in the vaccine (vaccine-serotypes) in diverse settings.(11,12)

Long term population-based IPD surveillance is ideal to monitor changes in the serotype distribution over time, with vaccination and in outbreaks.(13–16) IPD surveillance systems rely on comprehensive epidemiological, clinical and laboratory networks and are thus expensive to set up and maintain.(17) Unsurprisingly, IPD surveillance has been challenging in sSA, particularly population-based. The limited African data on serotype-specific IPD is dominated mainly by data from South Africa, with a few other countries reporting population-based IPD estimates and a few more, hospital-based estimates.(7,8,18–20) The few published studies that report serotype distribution for Nigeria are very old,(21,22) have very few isolates,(23–26) or are limited to specific IPD syndromes (meningitis),(27,28) and are hospital-based and not population-linked.

To compensate for the paucity of disease surveillance data, statistical models have been developed to estimate serotype distribution in IPD based on more readily available pneumococcal carriage data. The distribution of serotypes in IPD is determined, to a certain extent, by their distribution in nasopharyngeal carriage.(5,29) These models relate NP carriage prevalence to measures of invasiveness to predict IPD burden.(30,31) In this study, we applied two statistical models that express IPD as a product of serotype-specific carriage prevalence and its invasiveness,(30,31) using observed carriage prevalence data from rural and urban Nigeria locations to predict the relative contributions of serotypes to IPD prior to PCV10 introduction.

## METHODS

### Sources of data

#### Carriage prevalence

Nigeria introduced PCV10 to the childhood immunization program in phases between 2014 and 2016. We used carriage data from carriage surveys done in two sites at the time of PCV10 introduction (2016) in Nigeria. The surveys have been previously described.(32,33) In brief, the surveys were conducted among healthy residents of all ages in study sites in Ifo and Ado-Odo-Ota Local Government Areas (LGA, urban) and Kumbotso LGA (rural), southwest and northwest Nigeria, respectively. These sites are located in states in the last phase of PCV10 roll-out. For this pre-vaccine period, overall pneumococcal carriage prevalence was 92% in the rural site and, 71% and 78% in the two surveys in the urban site.(32,33) We extracted numbers of isolates for each serotype for U5s.

#### Invasiveness estimates

We used three measures of serotype invasive capacity (IC). The first estimated serotype-specific IC from a regression model using serotype case-fatality ratios from adult Danish IPD patients as predictors of serotype invasiveness.(30) Invasiveness was predicted for 37 serotypes.

The second approach estimated serotype IC as case-carrier ratios using NP carriage and IPD data from children in Kilifi, Kenya. We extracted NP carriage data of U5s from NP carriage surveys conducted before PCV10 introduction in the Kilifi Health and Demographic Health Surveillance Site (KDHSS)(34) We also extracted corresponding IPD data for KDHSS U5 residents admitted to Kilifi County Hospital (KCH). In this approach, IC for each serotype was estimated as follows:

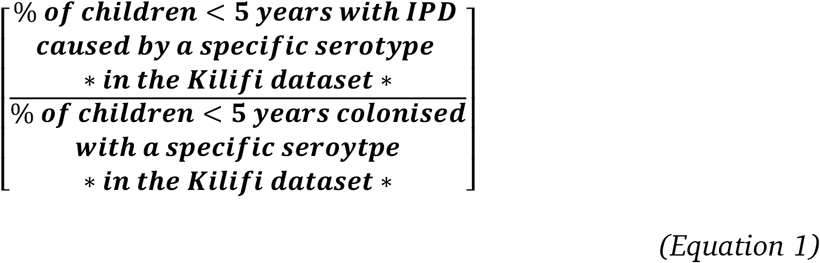

The third approach calculated serotype IC as serotype-specific attack rates using carriage acquisition incidence and IPD incidence in children in Oxford, UK.(35) In brief, longitudinal studies collected NP swabs at 4-weekly intervals from 2 to 96 weeks of age from a yearly birth cohort of 100 children between 1996 and 1999;(35) and another between 1999 and 2001 from a birth cohort of 213 swabbed at intervals, between 2 and 24 weeks of age.(35,36)The authors also collected IPD data covering 1995 to 1997 from active IPD surveillance of the same region from the National surveillance.(35,37) Serotype-specific attack rate was estimated as the ratio of IPD incidence to the incidence of NP acquisition.(35)

### Analyses

The first model, Model A, utilizes the serotype-specific log-carriage prevalence and log-estimated IC in a log-linked negative binomial regression to predict the number of expected IPD cases for each serotype.(30) The IC used in this model is serotype-specific case-fatality ratios from Danish adults, described earlier.(30)

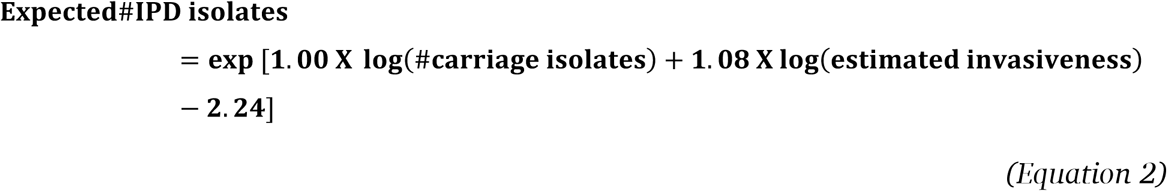

We used this model to predict IPD cases for the 37 serotypes for which the authors estimated invasiveness. We assigned an absolute value of 0.5 prior to log-transformation of carriage data to serotypes not detected in carriage.(30)

The second model utilizes serotype-specific carriage prevalence and IC.(31) Although, this model was originally used to model the decline in pneumococcal acute otitis media, it was validated using IPD data.(31) This model initially used carriage data in children aged <3 years from the US and estimated serotype-specific invasive capacity using data from children in Israel.(31) We ran this model twice using two different measures of serotype IC. In the first run (Model B), we used case-carrier ratios calculated from children in Kilifi, Kenya as measures of IC (34) to estimate the proportion of children colonized with a specific serotype expected to develop IPD. In the second run (Model C), we used attack rates from Oxford, UK(35) to estimate expected proportions of serotypes in predicted IPD. For serotypes not observed in carriage, we assigned an absolute value of 0.5 was. For serotypes where carriage was observed in the Kilifi or Oxford datasets, we imputed IC by assigning an absolute value of 0.5 to IPD isolates and consequently calculating case-carrier ratios or attack rates, respectively.

We grouped serotypes by different PCV types (PCV10, PCV13 and SII-PCV) to assess their relative contribution as a group to observed carriage and predicted IPD. The serotypes included in each PCV are shown in Table 1.

**Table 1:** Serotypes included in the different PCVs

| **PCV** | **Serotypes** |
| --- | --- |
| PCV10 | 1, 4, 5, 6B, 7F, 9V, 14, 18C, 19F, 23F |
| PCV13 | 1, 3, 4, 5, 6A, 6B, 7F, 9V, 14, 18C, 19A, 19F, 23F |
| SII-PCV | 1, 5, 6A, 6B, 7F, 9V, 14, 19A, 19F, 23F |

We did separate analyses for the urban and rural sites. Analyses were done in Stata 15.1 (Stata Corp LP, College Station, Texas).

### Model validation

To demonstrate the ability of the models to predict the relative contributions of each serotype as well as the proportion of IPD attributable to PCV10, we used carriage and IPD data available from Kilifi, Kenya. We applied Models A and C to observed Kilifi carriage data to predict IPD and compared these predictions to observed IPD from the same population using Spearman rank correlations.

## RESULTS

### Serotype-specific carriage

For this period, overall pneumococcal carriage prevalence in children aged <5 years was 92% in the rural site and 78% in the urban site, while carriage prevalence of PCV10 serotypes (VT) was 42% and 38%, respectively.(33) The four most prevalent serotypes were 6B, 19F, 23F and 6A accounting, collectively, for ≥50% of carriage at each site.

### Predicted serotype distribution in IPD

Model A predicted serotypes with high carriage prevalence (6A, 6B, 19F and 23F) to be the most common in IPD (Fig. 1A/B). The top five serotypes predicted from this model were 6A, 23F, 19F, 6B and 4 for the rural site, and 6B, 6A, 19F, 23F and 14 for the urban site. Models B and C predicted serotypes with high carriage prevalence and/or high IC estimates would predominate in IPD. The top serotypes predicted from Model B output were 4, 6A, 23F, 14, and 1 for the rural site, and 6A, 19A, 23F, 1, and 14 for the urban site. The top serotypes predicted from Model C output were 14, 4, 1, 6A, and 19F for the rural site and 14, 1, 19A, 19F, and 6A for the urban site.

**Figure 1A:**
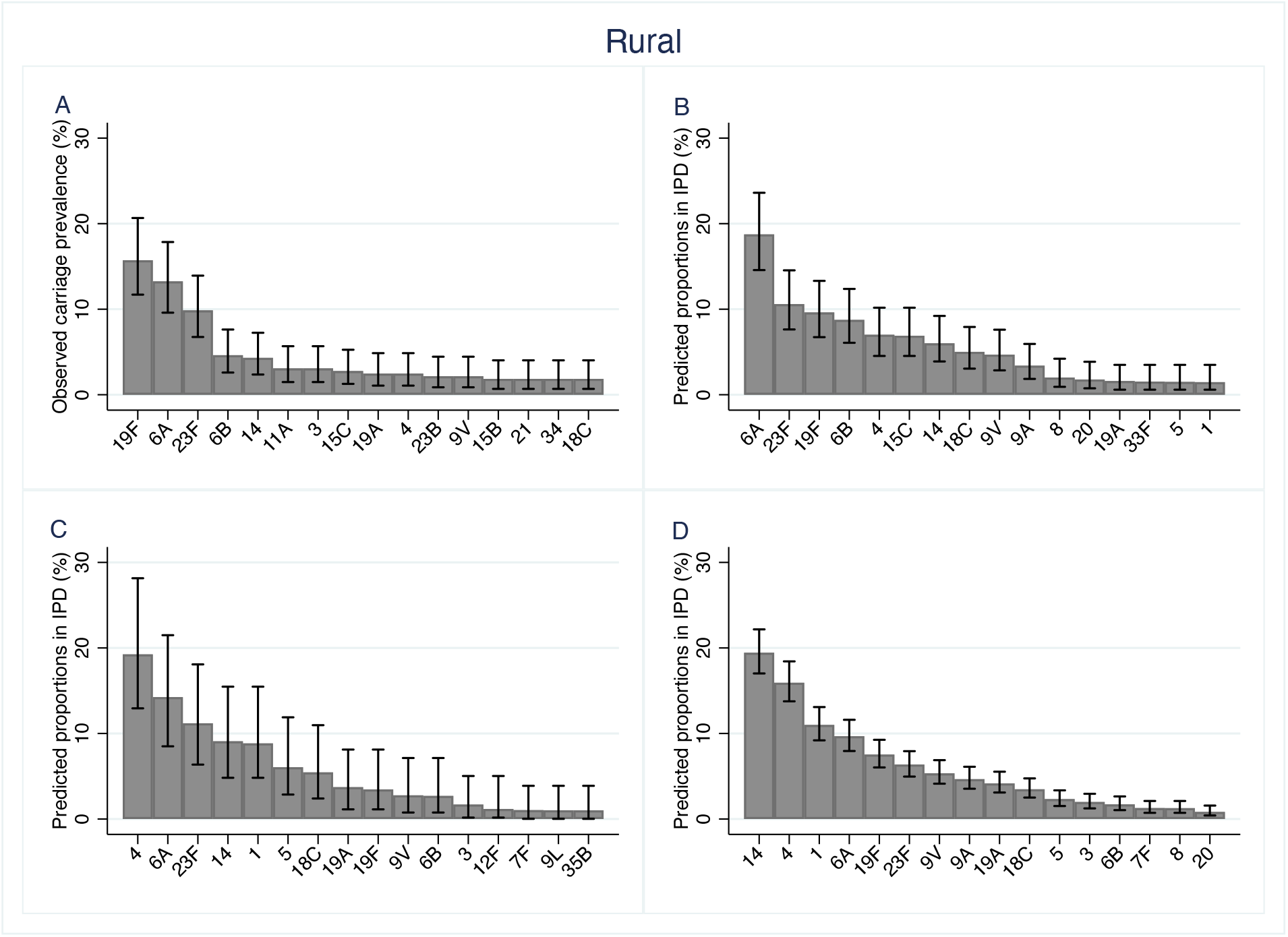
Observed serotype-specific carriage prevalence (A) and outputs from Model A (B), Model B (C) and Model C (D) showing predicted proportions of serotypes in IPD in the rural site

**Figure 1B:**
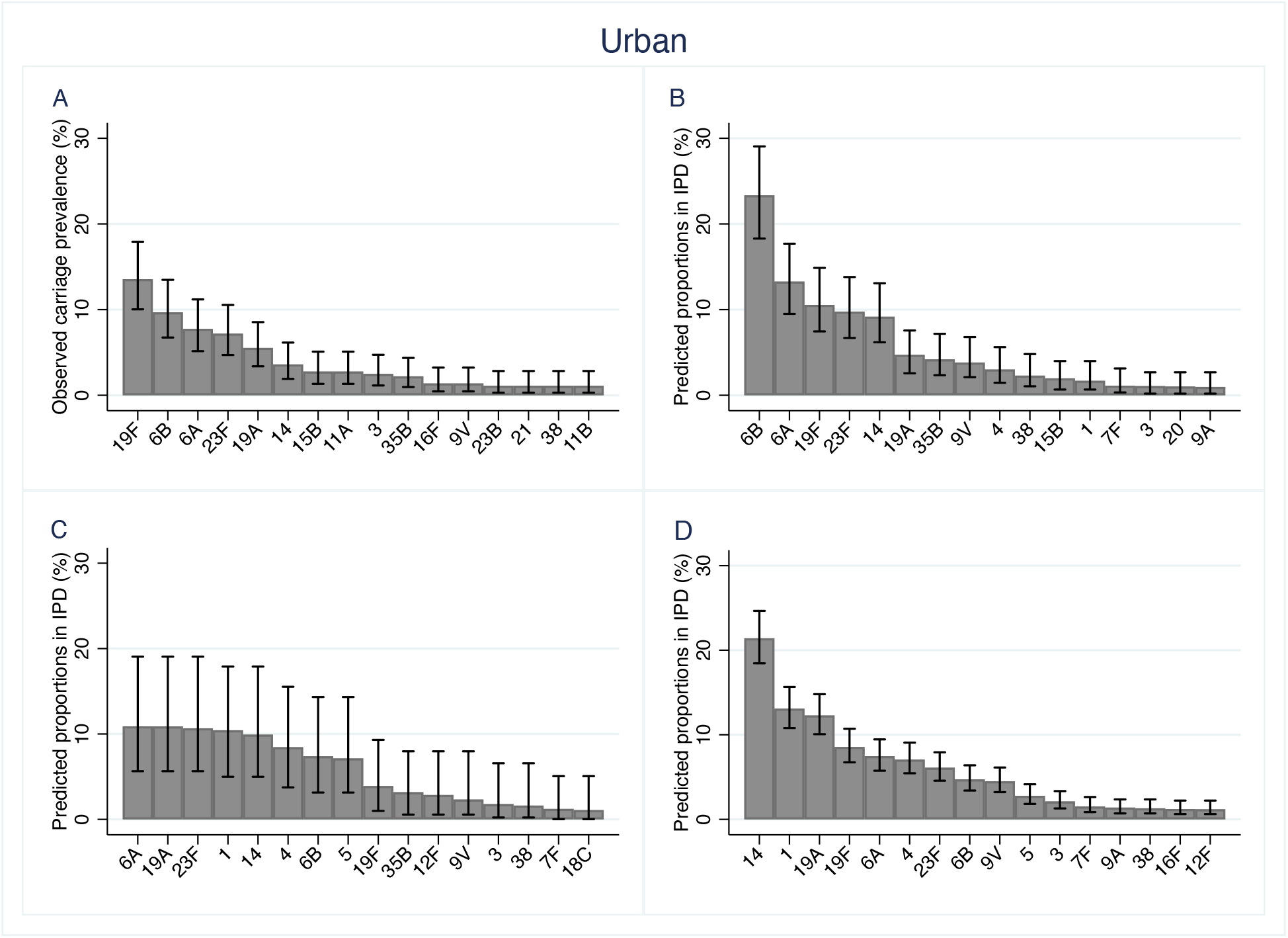
Observed serotype-specific carriage prevalence (A) and outputs from Model A (B), Model B (C) and Model C (D) showing predicted proportions of serotypes in IPD in the urban site

**Figure 2:**
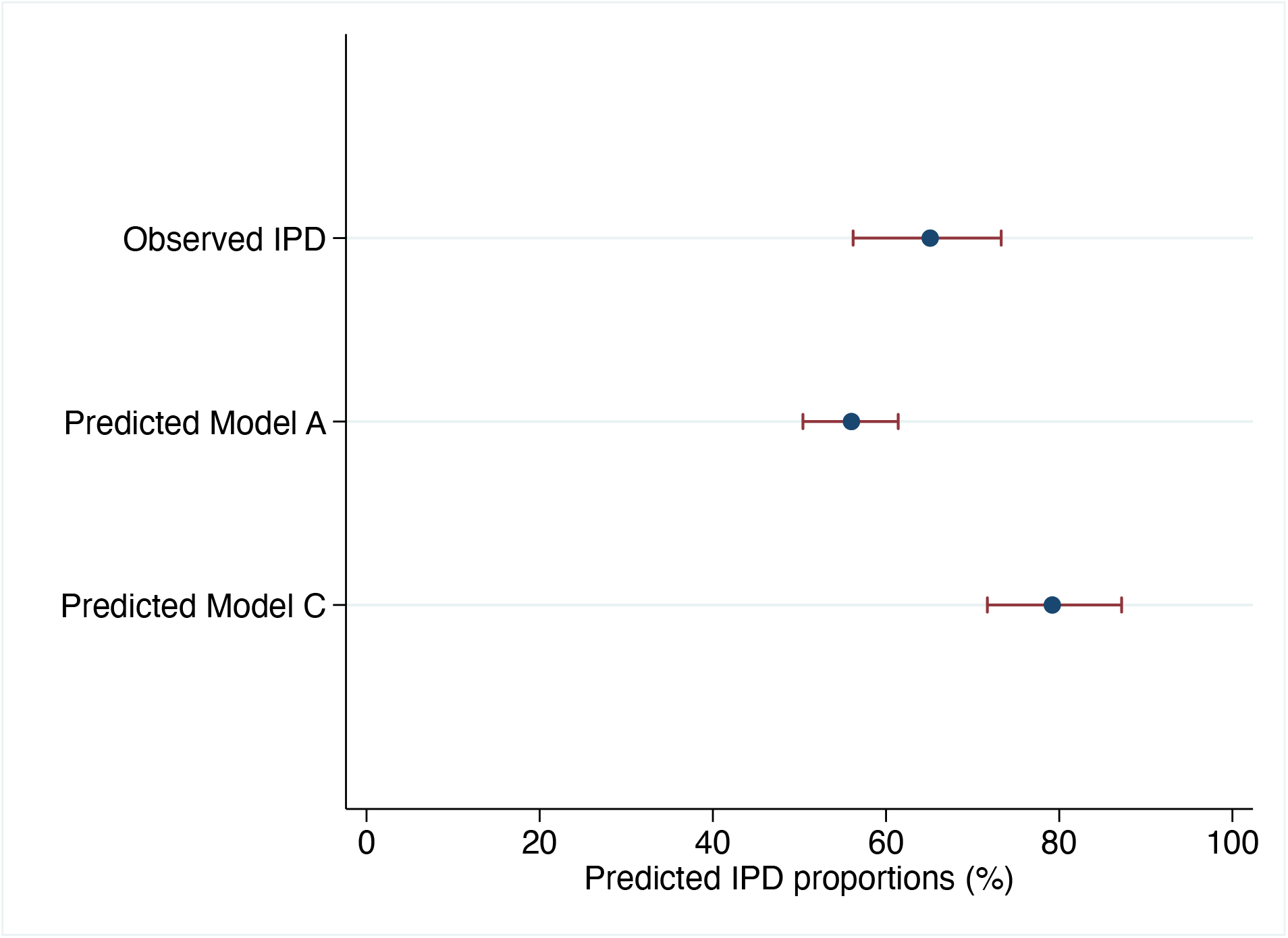
Relative contributions of PCV10 serotypes (grouped) in observed and predicted IPD in Kilifi, Kenya

### IPD attributable to PCV serotypes

Table 2 shows the proportions of serotypes covered by PCV10, PCV13 and SII-PCV in observed carriage and predicted IPD from the models. PCV10 serotypes accounted for 42% and 38% of all carriage isolates in the rural and urban sites, respectively. In both locations, carriage prevalence of PCV13 and SII-PCV were higher than PCV10. Model A output showed that PCV10 serotypes account for 56% and 64% of predicted IPD in the rural and urban sites. Outputs for Models B and C predicted higher proportions of IPD attributable to PCV10 serotypes compared to Model A in the rural site. In the urban site, the predicted proportions for PCV10 serotypes were similar across the models, but slightly higher for Model B (70%) compared to Model A (64%) or Model C (63%).

**Table 2:**
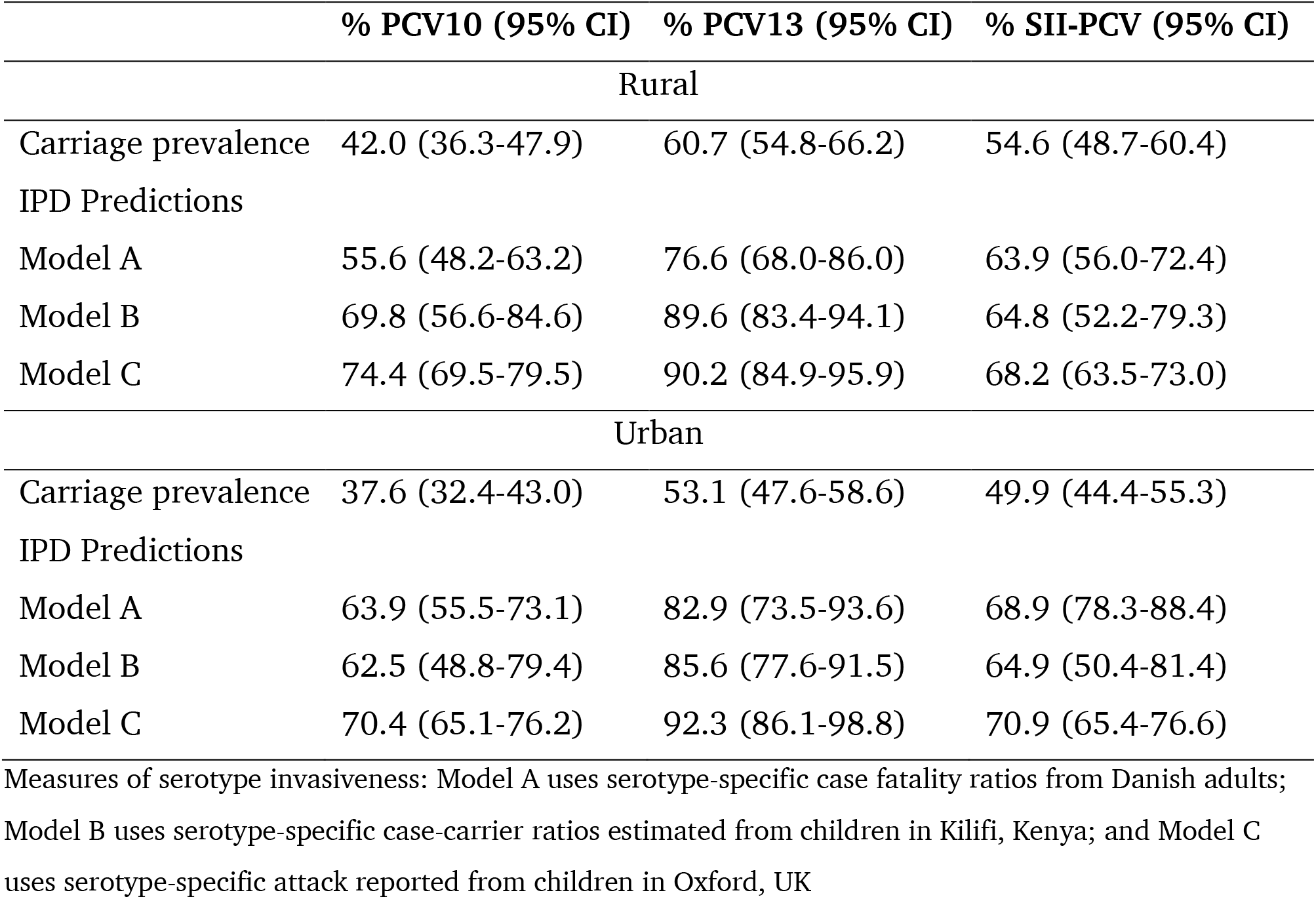
Relative contribution of serotypes in PCV10 in observed carriage and predicted IPD in the rural and urban sites

Model predictions for IPD attributable to PCV13 were higher than those of PCV10 or SII-PCV. Model predictions for IPD attributable to SII-PCV serotypes were similar to those of PCV10 across all three model outputs in both locations.

### Model validation

To demonstrate the performance of the models with observed data, we applied the model to carriage data from children in Kilifi, Kenya and compared the IPD predictions to observed IPD data from the same setting. We found a strong correlation between observed and predicted IPD in Kilifi for Model A (Spearman ρ=0.78, 95%CI 0.62-0.88), and Model C (Spearman ρ=0.81, 95%CI 0.65-0.90).

We also compared the proportion of IPD attributable to PCV10 serotypes in both the model predictions and the observed IPD data in Kilifi. Model A underestimated the proportion of IPD attributable to PCV10 serotypes, although the confidence intervals were overlapping, as shown in Figure 5. These differences were largely driven by serotype 1 which was underestimated significantly by the model and to a lesser extent by serotype 6B which was overestimated. For instance, the observed and predicted proportions of serotype 1 were 31.8% (95% CI:23.9-40.6%) and 6.7% (95% CI:4.3-10.0%), respectively; and for serotype 6B they were 8.5% (95% CI:4.3-14.7%) and 21.4% (95% CI:17.1-26.3%), respectively. Model C predicted a similar ranking of serotypes to the observed IPD data except for serotype 19F which was predicted in rank 3 but was observed in rank 8.

## DISCUSSION

Prior to widespread use of PCV, a small number of serotypes were responsible for a relatively substantial burden of IPD albeit with regional differences in the ranking of the serotypes. However, available data were largely dominated by sources from countries in North America and Europe and there was little data from sSA.(7,8,38) Accordingly, the first vaccine, PCV7, included serotypes that better represented the pattern of invasive disease in North America and Europe rather than sSA.(7) Defining the serotype distribution of IPD requires robust IPD surveillance but this is largely non-existent for many countries in sSA and what little data is available on serotype distribution comes from only a handful of countries.(19) In this analysis, we aimed to predict the distribution of serotypes in IPD among children aged <5 years in Nigeria using observed carriage data(32,33) and serotype invasiveness measures from previous models.(30,31) These models had previously replicated a multiplicative relationship between serotype carriage and invasiveness to predict serotype distribution in IPD and also replicated the observed burden of IPD in Kilifi, Kenya. Using these models, we predicted the serotype distribution and ranking in IPD prior to PCV10 introduction in Nigeria. We found similarities in the serotypes predicted to predominate in IPD from the models but with differences in the relative ranking of the serotypes.

The predicted proportions attributable to PCV13 were higher than either PCV10 or SII-PCV because of differences in serotypes valency and content. Although this may indicate that PCV13 may provide better protection, empirical data has shown that PCV13 has little to no protection against serotype 3 and has similar levels of serotype replacement as PCV10.(39,40) This higher predictions in serotype coverage for PCV13 may therefore not translate to larger IPD reductions. Our predictions show that SII-PCV has similar coverage to PCV10, and we attribute this to differences in carriage prevalence and IC of the included and excluded serotypes. Thus, any gains in coverage attributable to serotype 6A and 19A may be offset by non-coverage of serotype 4 by SII-PCV due to its relative high IC. The models predict that 13-22% of IPD are attributable to serotypes 6A and 19A. Thus, due to high predictions coupled with high carriage prevalence of serotypes 6A and 19A in our settings and lower vaccine cost, SII-PCV may have as or more public health impact than PCV10.(41)

The models predicted that serotype 1 accounted for ∼3% (Model A) and ∼12% (Model B and C) of all IPD. This is lower than what was previously reported for IPD by a hospital-based study from northern Nigeria where serotype 1 caused 31% (overall) and 48% (in children <12 years) of different IPD syndromes.(21) Even when broken down by IPD syndromes, this study showed higher proportions of serotype 1 for meningitis (45%), pneumonia (29%) and bacteremia (20%) than our predictions.(21) However, comparison to our model outputs may be challenging because this study was conducted between 1978 and 1980 when many serotypes were yet to be identified, and was hospital-based, with a substantial number of patients with co-morbidities that could additionally influence serotype distribution.(3) Moreover, the study did not present separate results for children <5 years and hence may not capture age-related differences in serotype distribution.(3)

Between 26-44% of IPD predicted by our models were attributable to non-PCV10 serotypes. Despite not having local IPD data to corroborate their distribution, these non-PCV10 serotypes are of clinical concern for a number of reasons. First, carriage frequency and duration provide a temporal opportunity for invasion regardless of invasiveness of a serotype especially with regards non-PCV serotypes 6A, 19A, 35B and 11A.(4) Second, although PCV10 provides variable cross-protection against 6A and to a less extent 19A, this protection may not be sufficient to reduce their burden in IPD, given their huge contribution to carriage in our context.(42) Lastly, these non-PCV serotypes already demonstrate a competitive advantage in carriage and thus may be important in serotype replacement in carriage, antimicrobial resistance, capsular switch, immune response evasion and serotype replacement disease.(43–45)

Although high levels of carriage and presumed poor acquired immunity in young children, can make carriage an important contributory factor to IPD burden in our context,(46) as predicted by Model A, we show that Model C had better predictions of more invasive serotypes (1, 4 and 5) as well as burden of PCV10-preventable IPD as validated by Kenya data. This indicates Model C had more accurate predictions compared to Model A. This can be explained by many reasons. IC estimates used in Model A may not be as appropriate because the authors only found a moderate correlation (0.58) between the adult case fatality ratios and actual invasiveness estimates (serotype-specific attack rates).(30) Additionally, case fatality may be a better measure of severity or virulence rather than invasiveness and therefore can be influenced by other host-or environmental-related factors. Moreover, serotype 1 reportedly has lower case-fatality rate in adults.(3,47) The IC estimates for Models B and C may be a closer measure of how probable it is for a serotype to cause invasive disease following a carriage episode. Both IC estimates used in this model were obtained from children (from Kenya or UK) and thus may be more accurate for predicting disease in children. From Models B and C, using case-carrier ratios and attack rates, respectively, we predict that at the time of PCV introduction, PCV10 serotypes would have contributed to between 70% and 74% of IPD in the rural site, and between 63% and 70% in the urban site. These values are similar to what was previously estimated for sSA (72%).(7)

The use of invasiveness and carriage to predict IPD is associated with some limitations. Invasiveness estimates are assumed to be stable and unique to serotypes, and this has been supported by some evidence.(48,49) However, factors that affect immune status can also influence the capacity of serotypes to cause invasive disease following carriage and these factors vary in different populations and may influence serotype distribution in IPD.(5) Bacterial factors unrelated to capsule can also affect serotype distribution in IPD. For instance, in The Gambia, an observed increase in serotype 1 disease in 1997 and 2002 was associated with the ST618 clone(15) and in Ghana, seasonal outbreaks of pneumococcal meningitis were associated with ST217 complex.(50) Therefore, the emergence of more virulent clones can increase the burden of some serotypes in IPD which will not be captured by the invasiveness estimates that rely on capsular serotype alone.

A second limitation is the absence of locally relevant IPD data in Nigeria to validate model predictions. Although we identified a few hospital-based studies in Nigeria that reported serotype distribution in IPD, these studies were either conducted a very long time ago(21,22) or had low culture yield and therefore few isolates.(23,24,28,51) However, using carriage data from Kilifi, Kenya, the model predicted the serotypes in IPD with only some slight differences in ranking between the observed and predicted. Thirdly, we could not predict ranking for all serotypes that were found in carriage because invasiveness estimates were unavailable for every serotype. However, given that carriage of these serotypes was low and invasiveness likely to also be low, it is reasonable to assume that these serotypes do not substantially contribute to IPD burden.

A portion of the carriage observations used in this analysis was not strictly pre-vaccine data because PCV10 had been introduced for four months prior to the carriage surveys in 2016. However, PCV10 was introduced only to infants who make up only a minority of the study population of children aged <5 years, and no catch-up vaccinations were given to older age groups. Furthermore, the ranking of the predicted serotypes did not differ when we limited our analysis to carriage data obtained in 2009 from the urban site.(32) Finally, a major assumption of our approach is that the IC is constant, within serotype, for each of the different IPD syndromes. Although prior evidence has shown differences in serotype ranking by IPD syndrome, we presume that IC will unlikely vary by IPD syndrome.(52)

In this analysis, we used serotype carriage prevalence and invasiveness estimates to model serotype distribution in IPD in Nigeria before PCV10 introduction. We predict that 56-74% of serotypes in IPD will be covered by PCV10, while SII-PCV serotypes will cover 64-71% of IPD. We demonstrate the value and limitations of using carriage data to predict serotype distributions in IPD in a typical sSA setting that lacks IPD data. SII-PCV may be a good alternative due to its lower cost and coverage of serotypes 6A and 19A that may indicate better protection against these prevalent serotypes. These predictions are a useful first tool to support vaccine policy in selecting and adapting the appropriate vaccine for use in Nigeria.

## Data Availability

Data supporting findings are included in the manuscript. Additional data requests can be made to the KEMRI-Wellcome Trust Research Programme Data Governance Committee

